# Patterns of Polysubstance use among Adults in Malaysia – A Latent Class Analysis

**DOI:** 10.1101/2022.02.14.22270961

**Authors:** R.H Wan Shakira, R. L Tania Gyle, S. G Shubash, A.M Nur Liana, MY Muhammad Fadhli

**Affiliations:** Institute for Public Health, National Institutes of Health, Ministry of Health Malaysia, Setia Alam, Selangor, Malaysia

**Author notes:** (WSRH).

**Keywords:** polysubstance use, illicit drug, alcohol, tobacco, kratom, Malaysia

## Abstract

**Introduction:** Polysubstance use is the use of more than one non-prescribed licit or illicit substance at one time. This is a common phenomenon, but little is known about the severity and the various substances used by adults in Malaysia.

**Objective:** To determine the pattern of polysubstance use and its associated factors among general adults in Malaysia.

**Methodology:** This was a secondary data analysis from the National Health and Morbidity Survey (NHMS) 2019), a cross-sectional population survey with a two-stage stratified random sampling design. A total of 10,472 Malaysians aged 18 years and above participated in this survey. Polysubstance use was defined as concurrent use of more than one substance, either alcohol, tobacco, or drugs (opioid, marijuana, amphetamine/ methamphetamine or kratom). A latent class analysis (LCA) was used to identify the membership of polysubstance groups. The association of class membership with demographic profiles was examined using Multinomial Logistic Regression analysis.

**Results:** Fit indices (AIC =16458.9, BIC = 16443.6) from LCA supported 3 classes solution: i) Combination of tobacco and alcohol (Tob+Alc) use (2.4%), ii) multi-drug use including kratom (0.3%) and iii) non/negligible user (97.3%). The multinomial model showed young adults (18-40 years) had a higher likelihood of being polysubstance users both for Tob+Alc class (OR=4.1) and multi-drug class (OR=3.9) compared to older age (≥60 years). Chinese (OR = 18.9), Indian (OR =23.3), Indigenous Sabah & Sarawak (OR =34.6) and others ethnicity (OR =8.9) showed higher odds of being Tob+Alc users than Malays. The greater odds of Tob+Alc. use for male (OR =35.5), working group (OR =1.5) and low education level group (OR=3.2).

**Conclusion:** Our study highlights patterns and demographics related to the use of polysubstance among adults in Malaysia. These results would help formulate specific prevention programmes for these high risk groups.

## Introduction

Polysubstance use is defined as the use of more than one non-prescribed licit or illicit substance. Licit substances include alcohol and cigarettes, while illicit substances include marijuana, cocaine, heroin, amphetamine, and methamphetamines [1]. In Malaysia, the most commonly used substances among adults were tobacco (21.3%), followed by alcohol (11.8%) and illicit drugs (0.5%), according to the findings of the National Health and Morbidity Survey (NHMS) 2019 [2]. Globally, the use of alcohol, tobacco and illicit substances are important contributors to the burden of morbidity and premature death [3]. Smoking had the highest substance-related death rate (110.7 deaths per 100 000 people), followed by alcohol and illicit drugs (33.0 and 6.9 deaths per 100 000 people, respectively) [4]. Although most of the studies focused on single substance use, many people especially illicit drug users, use more than one substance and were typically nested in a broader pattern of polysubstance use. Among the many possible combinations of polysubstance use, use of alcohol and other drugs combinations were the most common patterns [5,6]. However, in Southeast Asian countries, alcohol and tobacco was a common combination of polysubstance use [7].

Empirical research on various patterns of polysubstance use is important to identify the problem in the use of multiple substances. According to the National Epidemiologic Survey on Alcohol and Related Conditions (NESARC), most people with multiple substance use disorders (SUD) had at least one other co-occurring SUD. The prognosis of multiple SUD patients was worse than that of single SUD patients [8]. Furthermore, the synergistic interaction of multiple or polysubstance use has been shown to increase the possibility of negative consequences. Previous studies have pointed out that polysubstance use is associated with poisoning or overdose-related death [9-11], an increased risk of poor physical health, risky behaviour, poor response to treatment and mental health problems [12-14]. Epidemiological researches showed consistent links between polysubstance use and socio-demographic variables. Polysubstance use has been associated with young age [15], lower education [16] and socio-economic disadvantage [17]. Apart from that, some findings suggested that racial/ethnic differences in the pattern of polysubstance use [18,19].

In Malaysia, the pattern or class of polysubstance use in the adult population is still unclear. Despite having some information pertaining to polysubstance use in Malaysia, existing studies are limited to specific populations such as injection drug users [20,21], prisoners [22] and men who have sex with men (MSM) [23]. Therefore, it is necessary to investigate a nationally representative sample to provide necessary descriptive information for the development of theories explaining the pattern of polysubstance use. Moreover, the type of polysubstance use may vary in different countries. In Southeast Asia countries, kratom (the leaves from the tree *Mitragyna speciosa*) is one of the combined substances used by polysubstance users especially in Malaysia and Thailand. Kratom is an indigenous plant of Southeast Asia and it is reported to have similar effect on opioid and stimulants [24,25]. In Malaysia, substance users abuse kratom by mixing the kratom drink with other substances to obtain opioid-like effects or hallucinations. Singh et al (2014) reported that approximately 15% of the Malaysian population used Kratom to abstain from illicit drugs and alcohol [26].

Given the huge public health risk of substance use, understanding the pattern of polysubstance use should continue to be a priority. In this study, we used Latent Class Analysis (LCA) to identify patterns in the types of substances used (tobacco, alcohol, and other illicit substances) or latent class membership of polysubstance use. Currently, LCA is a method well used in substance use literature to explain the different patterns of substance use [27]. The purpose of this paper is to address the following research questions: (1) What are the class membership of polysubstance use and the prevalence of polysubstance use in each class membership in the general population? and (2) What kind of demographic profile can be used to predict the use of polysubstance in each class membership.

## Material and method

### Study design and sample characteristics

This study extracted sample data aged 18 years and above from the National Health and Morbidity Survey (NHMS) 2019. The NHMS 2019 focused on non-communicable diseases, their risk factors and other health problems, including substance use. This survey targeted residence in non-institutionalised living quarters (LQ) in Malaysia. NHMS 2019 is a cross-sectional population survey with a complex survey study design where the sample is representative of the entire Malaysian population. To ensure representativeness, this survey utilised a two-stage stratified random sampling. Population data from the Department of Statistics Malaysia (DOSM) was used as the sampling frame. The stratification involved all states in Malaysia including federal territories as the primary stratum. Within the primary stratum, urban and rural areas made up the secondary stratum. Sample selection started at Enumeration Blocks (EB) to the Living Quarters (LQ) and finally to the individual residing in the living quarters. The sample size was calculated using a single proportion formula to estimate prevalence with adjustment for; 1) finite population (based on 2019 projected population Malaysia), 2) design effect (based on previous NHMS 2015 survey), and 3) expected non-response rate of 35%. Thus, the optimum sample size required was 10,544 individuals above 18 years. Details of the sample size, sampling method and recruitment procedure have been reported elsewhere [2].

## Measures

### Measure of tobacco, alcohol, and drug use

The data for the alcohol and drug module were collected via a self-administered questionnaire while the tobacco module used an interview method.

### Tobacco

The questionnaire for tobacco was adapted from the short version of the Global Adult Tobacco Survey (GATS), which had been translated (into the Malay language), pre-tested and validated among the Malaysian population [2]. Current tobacco use was ascertained by the following question: “Do you currently smoke tobacco on a daily basis, less than daily or not at all?” Tobacco products included were manufactured cigarettes, hand-rolled cigarettes, kreteks, cigars, shisha, bidis or tobacco pipes. We defined current smoker as currently smoking any smoked tobacco products either daily or less than daily. This definition was based on GATS indicator guideline [28].

### Alcohol

Alcohol use was ascertained by the following questions: “In the last 12 months, did you consume any alcoholic beverage?” We defined current drinker as having consumed any alcoholic beverage in the past 12 months based on WHO’s definition of alcohol use [29].

### Drug use

The use of illicit or licit drugs was ascertained by the following questions: “During the past 30 days, did you use these types of drug/substance? 1) opioids (*heroin or morphine*), 2) amphetamines/methamphetamines, 3) marijuana, or 4) kratom”. We defined current drug use as taking or using any types of drugs opioids, amphetamines/ methamphetamines, marijuana or kratom in the past 30 days. The definition of drug use during the previous month (last 30 days) were well defined by the World Drug Report [30] and the European drug guidelines [31]

### Polysubstance use

Polysubstance use was defined as the current use of at least two psychoactive substances at the same time period, either combination of tobacco and alcohol, tobacco and drug, alcohol and drug or combination of all substances.

## Statistical Analysis

### Latent class analysis

The latent class analysis (LCA) approach was used to identify the pattern or group of polysubstance use. LCA is a statistical model used to explore the unobserved heterogeneity in a population and then assigning individuals into mutually exclusive and exhaustive types or latent classes based on their pattern of answers on a set of measured variables. It is a type of model-based cluster analysis that generally uses the expectation-maximisation (EM) algorithm for model estimation [32,33]. In our sample, we used six categorical substance variables as indicators for the latent class model, including current use of opioids (heroin/morphine), amphetamine/methamphetamine, marijuana, kratom, current smoker and current drinker. This LCA analysis was conducted using R software version 4.0.3 using the “poLCA” package [32]. We estimated a series of class models ranging from 2 to 5 classes (as we included only six indicator variables, not more than five classes were tested). We then evaluated the model to select the preferred model based on the following fit statistics: Akaike Information Criteria (AIC) and Bayesian Information Criteria (BIC). In general, lower AIC and BIC values indicate a better model fit as the lower value of the information criterion suggests a better balance between model fit and parsimony [27]. The Likelihood ratio and Chi-Square statistics also were used to assess model fit. Like the AIC and BIC the aim is to select models that minimise the Likelihood ratio and Chi-Square statistics whilst maintaining a low number of parameters. The larger the value of statistics, the more inefficient the model is to fit the data. After determining the best-fitting model, the posterior probabilities of group membership were used to assign participants to classes. The posterior probabilities refer to the probability of that observation that was classified in a given class.

### Multinomial regression

The multinomial regression analysis was used to assess the association between socio demographic characteristics with polysubstance groups based on LCA defined classes using STATA software version 15. In this study, a univariable analysis was carried out by testing all the 8 potential predictor variables (age, gender, ethnicity, strata, education, occupation, marital status, and household income) to screen for important independent variables. The variables with *p-*values <0.25 from univariable analysis were included in the preliminary final model (variable selection). The variable selection is the process of “reducing the model” to get the best fit model by including all the candidate variables in the model and repeatedly removing the variables with the highest non-significant p-value until the model contains only significant terms. Hence, the final model was created based on five variables significantly associated at the level of p <0.05 during the final steps of variable selection. Those variables were age, gender, ethnicity, education level and occupation status. Multicollinearity and interaction were checked accordingly. The overall fitness was checked using a Hosmer Lemeshow test, classification table and ROC (receiver operating characteristic) curve for each binary logit model. The findings were presented as crude and adjusted odds ratios with 95% confidence intervals.

### Ethical Approval

Prior to each interview, the purpose of the survey and methods used during the survey was explained to the respondent and information was handed out via the participant’s information sheet. Those who consented to participate were invited to answer the questionnaire module. This study obtained ethical approval from the Medical Research and Ethics Committee (MREC), Ministry of Health Malaysia (NMRR-18-3085-44207).

## Results

A total of 10,472 Malaysians above 18 years participated in this survey, giving an individual response rate based on the optimum sample required to be 99.3%. The demographic characteristics of survey respondents are shown in Table 1. The mean age was 32 years (SD = 3.7) with 43% of the sample aged between 18 to 40 years. The sample was predominantly female (54.3%), Malay ethnicity (64.5%), lives in urban areas (60.9%), secondary education level (47.7%), working (56.8%), married (68.3%) and had low household income (68.2%). Table 2 presented the weighted prevalence of substance use. Regardless of polysubstance use, the highest prevalence of single substance use was tobacco users (22.4%, 95%CI: 20.86, 23.96), followed by alcohol drinkers (11.8%, 95% CI: 10.04, 13.81) and drug users (0.5%, 95% CI: 0.37, 0.79). The total prevalence of polysubstance use was 4.6% (95%CI: 3.69, 5.78) where the combination of alcohol and tobacco (4.21%, 95% CI: 3.29, 5.37) was the highest prevalence among other possible combinations of polysubstance (Table 2).

**Table 1:** Socio demographic characteristic of participants (n= 10,472)

| Socio demographic characteristics | Count<sup>a</sup> | % |
| --- | --- | --- |
| Age (years), mean(±SD) | 31.7 (±3.7) | |
| 18-40 | 4502 | 43.0 |
| 41-59 | 3517 | 33.6 |
| 60 and above | 2453 | 23.4 |
| Gender |  |  |
| Male | 4785 | 45.7 |
| Female | 5687 | 54.3 |
| Ethnicity |  |  |
| Malay | 6751 | 64.5 |
| Chinese | 1327 | 12.7 |
| Indian | 662 | 6.3 |
| Indigenous Sabah & Sarawak | 1114 | 10.6 |
| Others | 618 | 5.9 |
| Strata |  |  |
| Urban | 6380 | 60.9 |
| Rural | 4092 | 39.1 |
| Education level |  |  |
| No formal education | 644 | 6.2 |
| Primary education | 2379 | 22.8 |
| Secondary education | 4969 | 47.7 |
| Tertiary education | 2425 | 23.3 |
| Occupation |  |  |
| Not working | 4520 | 43.2 |
| Working | 5944 | 56.8 |
| Marital status |  |  |
| Married | 7154 | 68.3 |
| Unmarried | 3318 | 31.7 |
| Monthly household gross income |  |  |
| Bottom 40% (B40) | 6702 | 68.2 |
| Middle 40% (M40) | 2325 | 23.7 |
| Top 20% (T20) | 795 | 8.1 |

<sup>a</sup>Unweighted count

**Table 2:** Prevalence of Substance use among adults in Malaysia aged 18 years and above (n= 10,472)

| **Types of Substance use** | **Prevalence (%) (95% CI)** |
| --- | --- |
| Current smoking | 22.40 (20.86, 23.96) |
| Current drinking | 11.80 (10.04, 13.81) |
| Current drug use (total)<sup>a</sup> | 0.50 (0.37, 0.79) |
| Heroin/morphine | 0.03 (0.01, 0.13) |
| Amphetamine or Methamphetamine | 0.10 (0.04, 0.22) |
| Marijuana | 0.10 (0.06, 0.29) |
| Kratom | 0.40 (0.22, 0.56) |
| Polysubstance (total)<sup>b</sup> | 4.60 (3.69, 5.78) |
| Alcohol + Tobacco | 4.21 (3.29, 5.37) |
| Alcohol + Drug | 0.05 (0.02, 0.12) |
| Tobacco + Drug | 0.43 (0.29, 0.64) |
| Tobacco + Alcohol + Drug | 0.04 (0.01, 0.10) |

Weighted prevalence (adjusted for design weight, non-response rate and population number)
<sup>a</sup> Total current drug use: used any type of drug in the past 30 days
<sup>b</sup> Total polysubstance used: having concurrent use of more than one substance at the same time period

### Latent class analysis

Results of model fitting for each model estimated from LCA are reported in Table 3. Based on these fit statistics, we selected a three-class solution as the best fit for the data. These three class solution presented the lowest AIC and BIC with the large improvement of likelihood ratio and chi-square Goodness of fit statistics values over the two class model. The three class solution added another class also did not appear to be meaningful and no significant improvement as the value for AIC and BIC increase after the three class solution.

**Table 3:** Latent class model fit statistics.

| Classes | AIC<sup>a</sup> | BIC<sup>b</sup> | Likelihood ratio | Chi-square goodness of fit | Maximum log-likelihood | Number of estimated parameter | df |
| --- | --- | --- | --- | --- | --- | --- | --- |
| 2 | 16458.86 | 16553.20 | 41.60 | 418.75 | -8216.432 | 13 | 50 |
| 3 | 16443.62 | 16550.75 | 12.36 | 24.13 | -8201.812 | 20 | 43 |
| 4 | 16453.77 | 16649.69 | 8.50 | 21.97 | -8199.885 | 27 | 36 |
| 5 | 16467.06 | 16713.78 | 7.79 | 13.39 | -8198.221 | 34 | 29 |

<sup>a</sup>AIC: Akaike Information Criteria, <sup>b</sup>BIC: Bayesian Information Criteria

The estimated class population share for the three class models is displayed in Figure 1. These are the estimated proportions corresponding to the share of observations belonging to each latent class [32]. The “Predicted class memberships” (posterior probabilities) is another way of estimating the size of the latent classes. Generally, when the values for the “estimated class population shares” and “Predicted class memberships” are similar, this is an indication of good model fit. The three class solution demonstrated almost similar values of estimated and predicted class solutions. Based on posterior probabilities, class 1, class 2 and class 3 contained 2.4%, 0.3% and 97.3% of the sample respectively. Class 1 and class 2 were classified as polysubstance group as they were characterised as having probabilities of using a combination of two or more substances at one period. Figure 2 present the probabilities of the different indicators in each class. The detailed probabilities for each indicator in the LCA model were described below;

**Figure 1:**
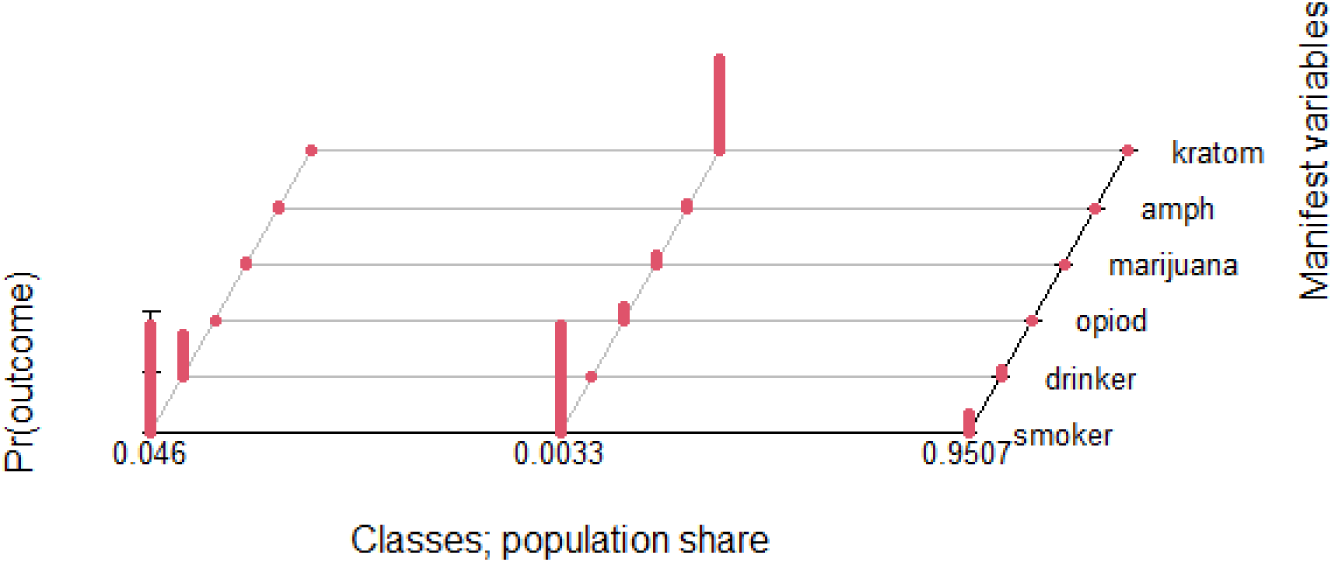
Estimates class population shares.

**Figure 2:**
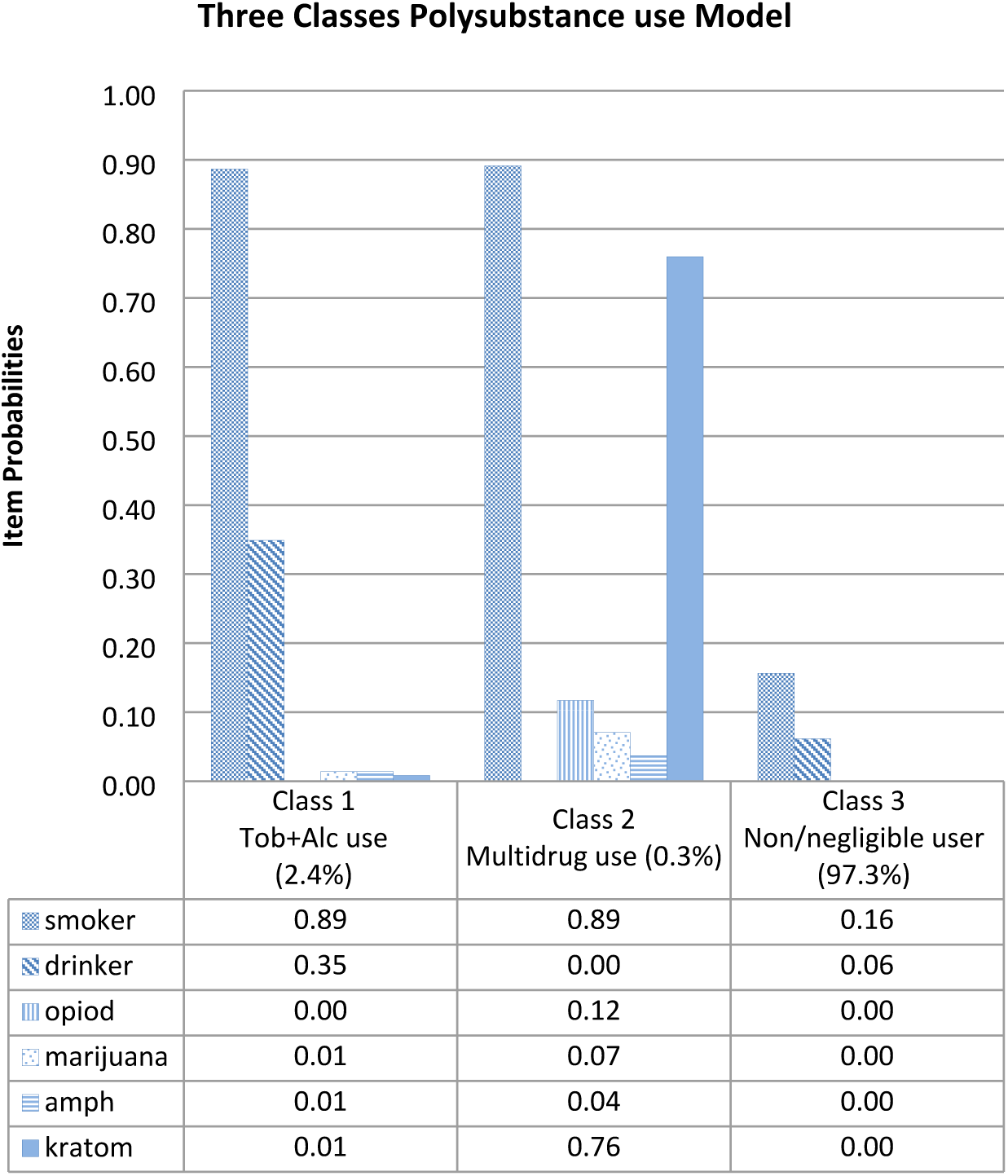
Probabilities of substance use indicators in each class in the three class solution.

- Class 1 was characterised by higher probabilities of tobacco and alcohol use compared to other classes, plus very low probabilities of all types of drug use. We named this class as combination of tobacco and alcohol use or “Tob+Alc use” class.
- Class 2 was the smallest sample (0.3%) but had high probabilities of smoking and kratom use with moderate to low probabilities of all types of illicit drugs, including opiods, marijuana, amphetamine/methamphetamines but no probabilities for alcohol use. This class named as “multi-drug use” class. Class 2 also demonstrated that our respondents had high probabilities of co-using of kratom with other illicit drugs.
- Class 3 contained the largest class (97.3% of the sample) characterised almost no probabilities of all types of substance except for tobacco and alcohol (low probabilities). We name this class as “non/negligible substance user” class.

The prevalence of each latent class membership by socio-demographic characteristics is presented in Table 4. Overall, majority of participants had negligible substance use or were non-substance users (class 3). The prevalence of polysubstance use according to latent class 1 (Tob+Alc) and class 2 (Multi-drug use) were 4.2% (95%CI: 3.25, 5.34) and 0.5% (95% CI: 0.31, 0.66) respectively. Young adults (age 18-40 years) demonstrated higher prevalence for both Tob+Alc used (5.1%) and multi-drug used (0.6%) compared with older adults. In terms of gender, males showed significantly higher prevalence of Tob+Alc use as compared to females. No prevalence of multi-substance use was estimated for females due to very low cases of multi-drug use among them. The combination of Tob+Alc showed high prevalence among the working group, primary education level, and non-Malay, especially Indigenous Sabah & Sarawak. However, Malays demonstrated high prevalence of multi-drug use (0.7%). The prevalence of polysubstance use for both Tob+Alc group and multi-drug group were relatively evenly similar among respondents from urban and rural areas, marital status groups and household income levels.

**Table 3:** Factors associated with polysubstance use among Malaysian Adults from Multivariate Multinomial Logistic Regression Analysis (n=10,464)

| Predictive factors | Univariate Multinomial Logistic Regression<br>Crude OR (95%CI) |  | Multivariate Multinomial Logistic Regression<br>Adjusted OR<sup>a</sup> (95%CI) |  |
| --- | --- | --- | --- | --- |
|  | Tob+Alc use<br>(Class 1) | Multi-drug use<br>(Class 2) | Tob+Alc use<br>(Class 1) | Multi-drug use<br>(Class 2) |
| **Age** |  |  |  |  |
| 18-40 years | 3.31 (2.14, 5.12)** | 3.25 (1.25, 8.40)* | 4.13 (2.45, 6.95)** | 3.85 (1.22, 12.12)* |
| 41-59 years | 2.15 (1.35, 3.42)* | 1.41 (0.48, 4.14) | 2.40 (1.41, 4.08)* | 1.60 (0.47, 5.44) |
| ≥60 years | 1 | 1 | 1 | 1 |
| **Gender** |  |  |  |  |
| Female | 1 | - | 1 | - |
| Male | 36.36 (17.95, 73.66)** |  | 35.51 (17.22, 73.25)** |  |
| **Locality** |  |  |  |  |
| Urban | 1 | 1 | - | - |
| Rural | 1.15 (0.88, 1.49) | 1.57 (0.87, 2.83) |  |  |
| **Ethnic** |  |  |  |  |
| Malay | 1 | 1 | 1 | 1 |
| Chinese | 14.05 (8.48, 23.29)** | 0.14 (0.02, 1.04) | 18.89 (11.16, 31.97)** | 0.18 (0.02, 1.30) |
| Indian | 19.44 (11.34, 33.33)** | 0.29 (0.04, 2.12) | 23.31 (13.30, 40.86)** | 0.32 (0.05, 2.32) |
| Indigenous Sabah&Sarawak | 30.52 (18.95, 49.14)** | 0.71 (0.35, 2.01) | 34.55 (21.02, 56.80)** | 0.76 (0.26, 2.16) |
| Others | 14.59 (8.20, 25.96)** | 0.31 (0.05, 2.24) | 8.88 (4.74, 16.62)** | 0.18 (0.02, 1.42) |
| **Marital Status** |  |  |  |  |
| Married | 1 | 1 | - | - |
| Single | 1.31 (1.01, 1.71)* | 1.37 (0.74, 2.51) |  |  |
| **Education** |  |  |  |  |
| No formal education | 1.83 (0.96, 3.49) | 0.42 (0.05, 3.33) | 3.23 (1.56, 6.68)* | 2.10 (0.23, 18.73) |
| Primary education | 2.10 (1.34, 3.29)* | 1.03 (0.41, 2.61) | 2.55 (1.56, 4.17)** | 2.25 (0.82, 6.19) |
| Secondary education | 2.29 (1.53, 3.44)** | 1.38 (0.64, 2.96) | 2.43 (1.58, 3.72)** | 1.48 (0.68, 3.19) |
| Tertiary education | 1 | 1 | 1 | 1 |
| **Occupation** |  |  |  |  |
| Not Working | 1 | 1 | 1 | 1 |
| Working | 4.14 (2.93, 5.88)** | 3.52 (1.64, 7.60)** | 1.50 (1.01, 2.25)* | 1.16 (0.50, 2.72) |
| **Income status** |  |  |  |  |
| B40% | 1.55 (0.89, 2.68) | 3.85 (0.52, 28.18) | - | - |
| M40% | 1.03 (0.56, 1.89) | 3.09 (0.39, 24.41) |  |  |
| T20% | 1 | 1 |  |  |

<sup>a</sup> Multivariate multinomial Logistic Regression was applied for adjusted odd ratio. Interactions were checked and no significant interaction term found. Overall fit the model for each binary logit was checked accordingly: Hosmer Lemeshow test (logit function 1, p = 0.842, logit function 2, p = 0.281), correctly classified table (logit function 1 = 99.03%, logit function 2 = 97.72%), Area under ROC curve (logit function 1 = 70.0%, logit function 2 = 92.5%).
\*p < 0.05, \*\*p < 0.001.

**Table 4:** Distribution of socio-demographic characteristics by latent class membership.

| Socio-demographic characteristics | Prevalence<sup>a</sup> (%) (95% CI) |  |  |
| --- | --- | --- | --- |
|  | Class 1<br>Tobacco+ Alcohol | Class 2<br>Multi-drug | Class 3<br>Non/negligible user |
| Total | 4.2 (3.25, 5.34) | 0.5 (0.31, 0.66) | 95.4 (94.22, 96.31) |
| Age (years) |  |  |  |
| 18-40 | 5.1 (3.73, 7.07) | 0.6 (0.39, 0.96) | 94.2 (92.35, 95.69) |
| 41-59 | 3.6 (2.48, 5.12) | 0.3 (0.16, 0.57) | 96.1 (94.58, 97.25) |
| 60 and above | 1.5 (0.88, 2.55) | 0.1 (0.05, 0.36) | 98.4 (97.32, 99.01) |
| Gender |  |  |  |
| Male | 7.9 (6.12, 10.07) | 0.9 (0.61, 1.28) | 91.2 (89.09, 93.02) |
| Female | 0.3 (0.12, 0.57) | - | 99.7 (99.43, 99.88) |
| Ethnicity |  |  |  |
| Malay | 0.5 (0.28, 0.93) | 0.7 (0.49, 1.07) | 98.8 (98.28, 99.12) |
| Chinese | 6.0 (3.80, 9.43) | 0.1 (0.01, 0.73) | 93.4 (90.46, 96.12) |
| Indian | 7.1 (4.45, 11.20) | - | 92.9 (88.77, 95.52) |
| Indigenous Sabah& Sarawak | 11.8 (9.03, 15.37) | 0.5 (0.14, 1.94) | 87.6 (84.22, 90.40) |
| Others | 8.4 (3.85, 17.38) | - | 91.6 (82.62, 96.14) |
| Strata |  |  |  |
| Urban | 3.8 (2.76, 5.27) | 0.4 (0.26, 0.68) | 95.8 (94.33, 96.83) |
| Rural | 5.4 (3.93, 7.42) | 0.6 (0.33, 0.98) | 94.0 (92.03, 95.54) |
| Education level |  |  |  |
| No formal education | 2.4 (1.29, 4.28) | 0.1 (0.02, 0.84) | 97.5 (95.59, 98.61) |
| Primary education | 6.9 (4.01, 11.55) | 0.3 (0.11, 0.59) | 92.9 (88.25, 95.75) |
| Secondary education | 4.7 (3.45, 6.39) | 0.6 (0.36, 0.97) | 94.7 (93.04, 95.98) |
| Tertiary education | 1.6 (0.95, 2.69) | 0.4 (0.21, 0.83) | 98.0 (96.89, 98.69) |
| Occupation |  |  |  |
| Not working | 1.5 (1.00, 2.30) | 0.2 (0.09, 0.46) | 98.3 (97.50, 98.82) |
| Working | 5.7 (4.37, 7.43) | 0.6 (0.39, 0.92) | 93.7 (91.99, 95.05) |
| Marital status |  |  |  |
| Married | 3.9 (2.85, 5.37) | 0.4 (0.24, 0.66) | 95.7 (94.24, 96.76) |
| Unmarried | 4.6 (3.08, 6.89) | 0.5 (0.32, 0.94) | 94.8 (92.60, 96.41) |
| Monthly household gross income |  |  |  |
| Bottom 40% (B40) | 4.9 (3.71, 6.36) | 0.5 (0.30, 0.72) | 94.7 (93.19, 95.84) |
| Middle 40% (M40) | 3.9 (2.12, 7.05) | 0.5 (0.24, 1.24) | 95.6 (92.48, 97.42) |
| Top 20% (T20) | 2.8 (1.57, 4.92) | 0.1 (0.01, 0.58) | 97.1 (94.99, 98.37) |

<sup>a</sup>Weighted prevalence (adjusted for: design weight, non-response rate and estimated number of population in Malaysian)

### Multinomial regression result

Table 5 presents the odds ratios (OR) comparing all other latent classes (polysubstance class 1 and class 2) to the “non/negligible user” class (class 3) as reference. In the fully adjusted model, gender, age group, ethnicity, education and working status were significantly associated with polysubstance class 1 (Tob+Alc). However, only age group was associated with polysubstance class 2 (multi-drug used). In summary, the younger age group (18-40 years) had higher odds of being polysubstance class Tob+Alc group [Adjusted OR (AOR) 4.13; 95% CI: 2.45, 6.95) and multi-drug group (AOR 3.85; 95% CI: 1.22, 12.12) compared with the older age group (60 years and above). Males had a higher likelihood of being Tob+Alc class users than females (AOR 35.51; 95% CI: 17.22, 73.25). Chinese ethnicity (AOR 18.89; 95% CI: 11.16, 31.97), Indian ethnicity (AOR 23.31; 95%CI: 13.30, 40.86), Indigenous Sabah & Sarawak (AOR 34.55; 95%CI: 21.02, 56.80) and other ethnicities (AOR 8.88; 95%CI: 4.74, 16.62) had a higher likelihood of being a member class of Tob+Alc as compared with Malay ethnicity. The odds of being Tob+Alc users were also higher among the working group (AOR: 1.50; 95% CI: 1.01, 2.25) versus the non-working group and low education level (no formal education, AOR= 3.23, primary education AOR = 2.55, secondary education AOR = 2.43) versus higher education level.

## Discussion

This study presents updated epidemiology of substance use data in the adult population of Malaysia. To our knowledge, this is the first paper to identify the pattern of substance use using LCA among a nationally representative adult population in Malaysia. We identified three distinct classes of substance use among the adult population: those who primarily use a combination of tobacco and alcohol (Tob+Alc), those who use multi-drug (including soft and hard drugs) and those who were negligible or non-drug users. This three class solution from LCA was interpretability and relevant fit statistics model. Interestingly, the latent classes we identified are somewhat consistent with the findings of previous work that investigated the profile of substance use among men who have sex with men (MSM) in Malaysia which identified a three class solution as the best fitting model: “negligible substance use”, “soft substance use” and “amphetamine-type stimulant use” [23]. Similar to our findings, they also found that majority of participants were negligible drug users or non-drug users and detected a small group of sample using hard drugs or multi-drugs.

Our work extends the literature in that we added kratom as one of the indicators for LCA in addition with other common substance use. We found that there was high probability of co-use of kratom with other multiple substance use (refer to class 2 latent class as visually represented in Figure 1 and Figure 2). This finding was supported by previous studies in Malaysia showed that substance users in Malaysia used kratom to abstain from illicit drugs and to manage stimulant withdrawal symptoms [25,26,34]. A study on kratom consumption in the northern areas of peninsular Malaysia also reported that they used kratom to reduce their intake of more expensive opiates like heroin [25]. Although kratom has perceived therapeutic effects, several studies suggested abuse and addiction potential, synergistic toxic effects and fatal interactions with other psychoactive drugs [35,36]. Furthermore, there has been no report on the fatalities in Southeast Asia caused by the ingestion of pure kratom alone. Although the enforcement of kratom related offences in Malaysia is under the jurisdiction of the Poisons Act 1952 [37], the issue of misuse and offences must be addressed by all parties including law, policymakers and the community to improve the enforcement work done by the authorities.

In our Multinomial regression results, we chose Class 3 (non/negligible substance use) as the reference category to distinguish the pattern of polysubstance use from predominantly negligible or non-users. As reported in numerous literature, gender (male) is an important demographic characteristic associated with polysubstance use [16,19,38]. This study detected a significant association of the male gender with the Tob+Alc group but not for the multi-drug group. This may be due to a very low prevalence of multi-drug use among female respondents. In terms of age differences, the young adults (18 – 40 years) demonstrated higher prevalence and risk of being a member of polysubstance use than older adults (≥60 years), for both Tob+Alc and multi-substance classes. Our findings are in line with the study from The United States that reported the co-use of tobacco and alcohol were highest among young adults and declined with increasing age [39]. As well as for other polysubstance the risk of use, abuse/dependence and use of other forms of illicit drugs after the use of marijuana/cannabis declined with increasing age [15]. Another LCA study among the Great Britain population also reported that being young was associated with an increased likelihood of membership of the polydrug use class’s wide range and moderate range [38]. There are several explanations of this trend for polysubstance use to decline with increasing age. This includes socio maturity where increasing age will increase the social maturity on the individual’s ability to resist other illicit drug use [40]. Other explanations might be due to a recruited effect or age of onset of first drug use such as marijuana, those who take up marijuana later would be less predisposed to other illicit drug use than those who take it up early.

Our findings of polysubstance use varying among racial/ethnic groups are consistent with the results from other studies [16,19,39,41]. For example, a study in the United States found that Whites were more likely to drink while Natives American/Alaskan Natives were most likely to smoke and co-use alcohol and tobacco [39]. Our results showed that the co-use of alcohol and tobacco were more likely to occur among Indigenous Sabah & Sarawak, followed by Indian and Chinese ethnicity as compared to Malay ethnicity. However, according to the LCA model, the Malays ethnicity demonstrated the highest prevalence of multi-substance use (which had low probabilities of taking alcohol) compared to other ethnic groups. Our study was in line with the findings noted in another epidemiological study in Malaysia reporting similar results, although that study involved adolescents [42]. These findings suggest that the culture and practices of respective ethnic groups might play an important role in predicting substance use. In Malaysia, Malay adults had low prevalence of alcohol use but high prevalence of tobacco use, while Indigenous Sabah & Sarawak showed high prevalence for both alcohol and tobacco consumption [2,43]. In this country, Muslims (the majority are of the Malay ethnicity) are forbidden from consuming alcohol, and it is also illegal to sell any alcoholic beverages to them. In contrast, drinking alcohol is a culture and social obligation among other ethnic groups.For example, indigenous Sabah & Sarawak consume alcohol during their festivals and social gatherings [44].

The present findings also suggested that adults with low education levels (no formal education, primary education and secondary education) were more likely to use polysubstance of Tob+Alc group compared to higher education levels (tertiary education). However, no significant association was found between education levels and polysubstance of multi-substance group. Our findings is consistent with the latent class analysis of polysubstance from European studies which reported higher education was associated with a reduced risk of moderate but not high polysubstance class [16,38]. They found that there was a reduced risk of moderate polysubstance group membership for participants with an education level beyond secondary education (General Certificate of Secondary Education, GCSE).

Our study also revealed that working adults is a significant predictor for polysubstance use in the Tob+Alc class. This is consistent with the epidemiological data in Malaysia, which reported that the prevalence of current smokers and drinkers were higher among working groups, especially self-employed and private employees compared to those not working [2,43]. Working conditions may influence addictive behaviours such as exposure to psychological job strain (due to high work demands and low coping skills) that may lead to an increased risk of substance use especially alcohol and tobacco [45-47]. The misuse of alcohol by workers represents an important social policy issue because it has the potential to influence the employee’s job performance. For instance, even moderate daily alcohol consumption can lead to absenteeism and smoke breaks among employees that can be considered disruptive as it takes time away from work.

Taken together, the results from the current study suggest that prevention and treatment strategies for polysubstance use especially for the co-use of alcohol and tobacco should place a special emphasis on young adults and other high-risk groups such as people from the Indigenous Sabah & Sarawak ethnicity and those who are working. Whilst, the prevention strategies for polysubstance use of multi-drugs including illicit drugs, should focus on young adults and people from the Malay ethnicity. In addition, kratom use should not be reasonably expected to be safe, especially for co-use with other drugs, and it poses a public health threat. Thus, stronger measures to control of this drug are warranted and the use of kratom in Malaysia should be further investigated, including the effects of co-use with other drugs.

This study needs to be interpreted with caution in light of several limitations. The primary limitation of this study is the relatively low prevalence rate of multi-drug uses such as opioids, marijuana and amphethamine type stimulants (ATS). Due to the small number of cases of illicit drug use, our study may have been underpowered to identify additional classes and differentiate the pattern of polysubstance classes between hard drugs and soft drugs in the multiple-drug group (class 2) and other potential variables associated with multi-drug use. Our study also used self-reported substance use where the prevalence rates could be misrepresented. However, past researchers suggested suitable reliability and validity of studies with self-reported substance use [48,49]. The limitation of this study was the sample recruited from a cross-sectional design of a population-based survey, and it only investigated the socio-demographic predictor factors related to polysubstance use. Although other studies [50-52] have found the association between polysubstance use and psychological distress, as our study extracted the data from a large population-based survey which is almost similar to other existing population-based studies mainly cross-sectional, we have no capacity to consider prospectively early life predictors and to describe the association with psychological disorder. Future studies should replicate and extend these findings with a larger sample using a cohort design and to measure the association of polysubstance use with other early life predictors and mental health disorders.

## Conclusion

In summary, the current study provides an illustration pattern of polysubstance use in Malaysia with a three class solution (Tob+Alc, Multi-drug and non/negligible user). The LCA model also demonstrated a high probability of co-use of kratom together with other illicit drugs in the multi-drug classes. Our study emphasises the importance of considering patterns of polysubstance use when addressing demographic risk factors. As noted in our regression model, there was an increased risk of being a multi-drug user among the younger age group. While the model for Tob+Alc class demonstrated a strong link with males, young adults, non-Malay ethnicity, working groups and low education level.

## Data Availability

The National Institute of Health, Ministry of Health Malaysia (data ethics committee) has placed restrictions on sharing the full dataset due to cases involving researchers manipulating the data. Interested researchers will need to send a formal letter/email to the Director General of Health Malaysia, together with the data request form and proposal, available at (http://iku. moh.gov.my/images/IKU/Document/Form/ Borangpermohonandatalatest.pdf). The proposal will be reviewed by the Data Repository team from Biostatistics Sector, National Institute of Health Malaysia to ensure no duplication with other projects that have used the NHMS 2017 data. The data request flow chart is available on the website and can be accessed here (http://iku.moh.gov.my/ images/IKU/Document/Form/ FlowChartforIPHDataApplication.pdf). The authors also, confirm they did not have any special access privileges that others would not have for the NHMS 2017 data. The contact information of the Director General of Health Malaysia as below Address: Director General?s Office, Ministry of Health Malaysia, Kompleks E, Aras 12, Blok E7, Presint 1, 62590 Putrajaya, Malaysia Email: anhisham@moh. gov.my Contact number: +60388832545, Fax number: +60388895542

## Acknowledgement

We would like to thank the Director-General of Health Malaysia for his permission to publish this article.

